# Number of children and mid- to later-life cognitive function and cognitive impairment in rural South Africa: Evidence from “Health and Aging in Africa: A Longitudinal Study of an INDEPTH Community in South Africa” (HAALSI)

**DOI:** 10.1101/2023.09.25.23296101

**Authors:** Meredith L Phillips, Lindsay C Kobayashi, Rishika Chakraborty, Ryan Wagner, Nomsa Mahlalela, Jaroslaw Harezlak, Joshua W Brown, Christina Ludema, Molly Rosenberg

## Abstract

**Background:** Cognitive impairment is projected to rise substantially by 2050, particularly in low- and middle-income countries with aging populations. Reproductive life history may be associated with later-life cognitive function. We aim to estimate the association between number of children and mid- to later-life cognitive performance.

**Method:** Data were from 5059 older adults (46% men) aged ≥40 years in the population-representative rural cohort of the “Health and Aging in Africa: A Longitudinal Study of an INDEPTH Community in South Africa” (HAALSI). We fit linear regression and modified Poisson models to estimate the associations between number of children and cognitive function. Analyses were stratified by sex/gender, controlling for age, education, literacy, self-reported childhood health, country of birth, and father’s occupation.

**Results:** After adjustment, men with any number of children and women with 5+ children had higher cognitive function compared to those without children (β[95% CI]: Men: 1-2: 0.29[0.13 – 0.45], 3-4: 0.50[0.34 – 0.65], 5+: 0.48[0.33 – 0.63]; Women: 5+: 0.17[0.01 – 0.34]). Results from the adjusted modified Poisson regression models found that for men and women, groups with any number of children showed a lower prevalence of cognitive impairment than the group with 0 children (PR[95% CI] Men: 1-2: 0.48[0.31 – 0.75], 3-4: 0.21[0.12 – 0.37], 5+: 0.28[0.19 – 0.41], Women: 1-2: 0.52[0.31 – 0.88], 3-4: 0.55[0.33 – 0.91], 5+: 0.41[0.25 – 0.69]).

**Conclusion:** Among older rural South African adults, having children was associated with greater cognitive performance and lower prevalence of cognitive impairment. Men tended to have larger protective associations than women, which may be due to sex/gender differences in biological and social roles of childbearing and rearing. Understanding the complex relationship between having children and later-life cognitive function may help identify interventions to reduce the impact of cognitive decline.

## INTRODUCTION

The global prevalence of accelerated cognitive decline and impairment is projected to rise rapidly in the coming decades, especially in low and middle-income countries where life expectancy is increasing.(1–3) Identifying modifiable risk factors for aging-related cognitive impairment is thus an important global public health objective. Modifiable risk factors for cognitive impairment include years of formal education; mental health conditions, such as depression; cardiovascular risk factors, such as insulin resistance and hyperlipidemia; lifestyle risk factors, such as cognitive training and social engagement; and sleep disturbances.(3–5) Some of these risk factors can be linked to the social and biological experiences of parenthood, meaning that reproductive life history, including number of children, may be associated with later-life cognitive function, decline, and impairment,(6–20) It is essential to better understand the potentially complex relationship between parity and later-life cognitive health outcomes, to identify potential interventions to reduce the population health impact of cognitive decline.

During the parenting experience, different risk and protective factors could plausibly influence cognitive function. These experiences begin at conception for women and last throughout their lifespan. While men are not exposed to the physical demands of carrying a pregnancy, they can still experience the demands and benefits of childrearing. Physiologically, mothers and fathers experience changes in hormones and other metabolic processes, which can persist into later life and contribute to cognitive performance.(6–11) Mothers can also experience cognitive deficits that begin in pregnancy and can last years following parturition.(12) Parents often experience increased sleep deficits and disturbances compared to nonparents.(13,14) Being a parent can increase lifetime chronic stress, but it can also increase social support and cognitive stimulation.(15–20) The sum of all the reproductive life history experiences could plausibly influence cognitive performance positively or negatively for mothers and fathers in mid- to later-life (Figure 1).

**Figure 1.**
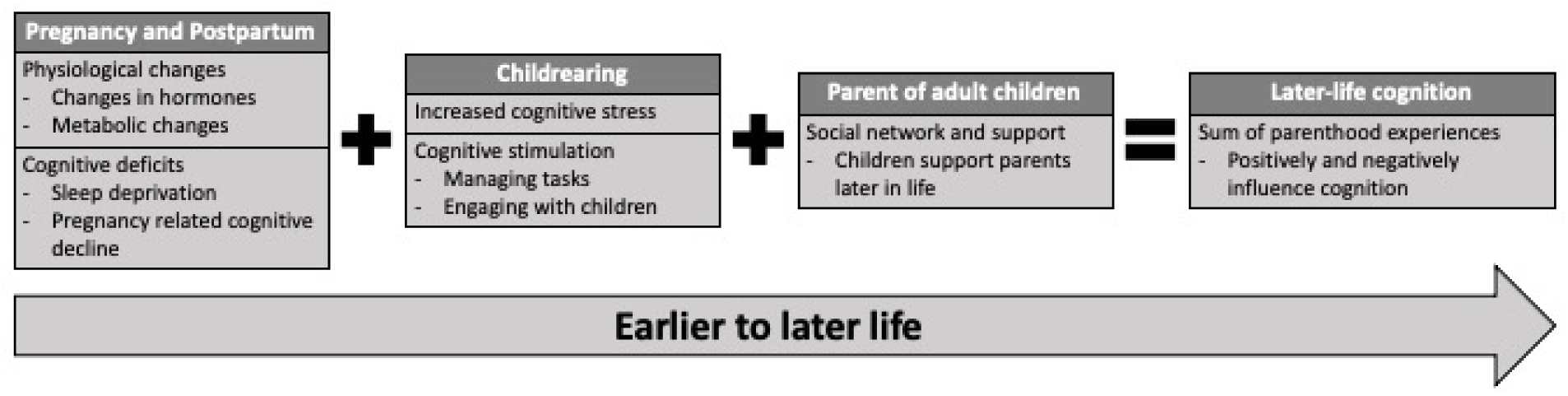
Conceptual model

One way to measure reproductive life history is by looking at the number of children someone has, which may be associated with mid- to later-life cognitive performance.(21) For women, prior work has suggested that the relationship between number of children and cognitive performance is an inverse U-shape,(22–26) with lower cognitive performance at a low and high number of children. Other work has suggested that it is a negative relationship, with an increasing number of children being associated with lower cognitive performance.(27–37) For men, the association is also unclear, with a small number of studies finding either a U-shaped relationship,(23–26) a negative relationship,(28) or no relationship at all. (34,38) Further examination of this relationship is essential to support the identification of potential intervention targets, including educational opportunities around family planning or childrearing or provision of other resources for parental support that target the negative impacts of childrearing, which are critical in low- and middle-income countries with rapidly increasing cognitive impairment prevalence and incidence.

With this study, we aimed to determine the association between the number of living children and mid to later-life cognition by sex/gender. We hypothesize that the sum of the child-birthing and rearing experiences throughout a person’s life will impact their cognitive aging, positively impacting cognition. Using data from the population-representative cohort in a rural South African community, “Health and Aging in Africa: A Longitudinal Study of an INDEPTH Community in South Africa” (HAALSI), we estimated the association between number of living children and mid- to later-life cognitive function and cognitive impairment.

## METHODS

### Study population

We used data from baseline interviews with a cohort of men and women aged ≥40 years in the population-representative “Health and Aging in Africa: A Longitudinal Study of an INDEPTH Community in South Africa” (HAALSI) in a rural Agincourt sub-district, Mpumalanga province, South Africa.(39) The study area is located near the border of Mozambique and houses a high proportion of Mozambican refugees.(39,40) The Agincourt area was used during the Apartheid era as a resettlement location for black South Africans, resulting in forced racial segregation.(40) After the Apartheid era ended in 1994, economic and educational opportunities in the area somewhat improved. However, the area remains low-income, with high unemployment rates and an older population with severely limited access to education during their youth.(40) Additionally, fertility in this population has fallen over the past years; the average total fertility rate was 6.0 in 1979, fell to 2.3 in 2004, and remained relatively stable.(41,42)

HAALSI is a harmonized International Partner Study to the US Health and Retirement Study (HRS) and other global aging cohorts.(39) HAALSI was designed and implemented with the aim of better understanding how aging in a rural South African population affects cognitive and physical health. The HAALSI sample was drawn from the Agincourt Health and Demographic Surveillance System (HDSS), which collects annual census data from approximately 116,000 people residing in 31 contiguous villages (39,40). Eligible participants for HAALSI were randomly sampled from all permanent residents living in the study setting who were aged ≥40 as of July 1, 2014. Of the 6,281 individuals randomly selected for recruitment in HAALSI, 5,059 participants (2,345 men and 2,714 women [53.6%]) provided written, informed consent and completed in-home interviews (86% response rate).

### Patient and Public Involvement

Patients or the public were not involved in the design, conduct, reporting, or dissemination of our secondary data analysis.

### Data collection

Trained fieldworkers visited study participants in their homes and collected baseline data from 2014-2015 using a Computer Assisted Personal Interviews (CAPI) platform in the local xiTsonga language. These interviews covered household and individual-level questionnaires, clinical data, such as anthropomorphic measures and biomarkers, cognitive assessments, and demographic information.(39) Data is publicly available.(43)

### Key variables

#### Exposure variable

We computed our exposure of interest, number of living children, by first assessing the response to the question “Do you have any children?” with response options being ‘yes’ or ‘no.’ If a participant answered ‘no,’ we entered them as having ‘0’ children. If participants answered ‘yes,’ they were then asked, “How many living children do you have?” We used participants’ responses to this question and categorized them:(44–47) none (0 living children), low (1 to 2 living children), moderate (3 to 4 living children), and high (5 or more living children).

To check the robustness of our findings, we used two additional exposure variables in supplemental analyses. First, we summed the number of self-reported living children, as indicated above, with the number of self-reported deceased children and categorized them into the previously defined categories. Second, we used the continuous, rather than categorical, number of living children with a quadratic term to account for the potentially non-linear relationship between number of children and the outcome.

#### Outcome variables

We examined two outcomes: 1. cognitive function and 2. cognitive impairment. Cognitive function was assessed using a continuous z-standardized latent variable encompassing orientation (the ability to state the current day, month, year, and South African President), episodic memory (immediate and delayed recall of a 10-word list spoken out loud by the interviewer), and numeracy (the ability to count forwards from 1-20 and complete a number skip pattern).(48) If the participant could not complete the interview due to severe cognitive impairment, physical illness, or deafness, proxy interviews were conducted with a friend or family member. Cognitive impairment was classified as having a cognitive function z-score ≤1.5 standard deviations below the sample mean, or requiring a proxy interview with proxy-reported memory as “fair” or “poor”.(49)

#### Covariates

We examined a set of covariates to explore their role as confounders and effect measure modifiers: sex/gender (male or female), age in years, self-reported childhood health (very good, good, moderate, bad, and very bad), country of birth (South Africa or other), father’s occupation level during childhood(48,50) (Skill level 1 [unskilled manual labor], Skill level 2 [mining and service industry], Skill level 3 [traditional healers and assistants), Skill level 4 [professional], other), education (some formal education or no formal education), and self-reported literacy (ability to read or write: yes or no). Gender information was not collected separately from sex.

### Statistical analysis

We estimated the association between the categorical number of children and 1) cognitive function and 2) cognitive impairment. As the influence of having and raising children on later-life cognitive outcomes could plausibly vary by sex/gender, we stratified all analyses by sex/gender. We fit sex/gender-stratified multivariable linear regression models (cognitive function outcome) and modified Poisson regression models (cognitive impairment).(51) For all analyses, having 0 children was the reference category. Our first models were unadjusted, including no covariates. The second models included a limited adjustment set of known risk factors for cognitive decline (age, education, literacy). The third models included an adjustment set of covariates from model two and additional socioeconomic covariates associated with this population’s cognitive performance (age, education, literacy, self-reported childhood health, country of birth, and father’s occupation level during childhood). We compared models for best model fit using the Akaike information criterion (AIC). After model selection, we completed Tukey post hoc analyses to compare differences across the categorical number of children groups. Due to relatively low missingness (all <12%), we conducted a complete case analysis. We conducted all statistical analyses in RStudio.(52)

As sensitivity analyses, we fit the above models with the two additional exposures: 1) the categorical number of all children (living and deceased) for both outcomes and 2) the continuous number of children with a quadratic term for the cognitive function outcome only.

### Ethical considerations

Researchers obtained written, informed consent from all participants in the local language, xiTsonga, or English. If a participant was not literate, a witness was present, and the participant provided a fingerprint instead of a signature. Researchers obtained ethical approvals for the study from the University of the Witwatersrand Human Research Ethics Committee (Medical) (ref. M141159), the Harvard T.H. Chan School of Public Health, Office of Human Research Administration (ref. C13–1608–02), and the Mpumalanga Provincial Research and Ethics Committee. Indiana University Bloomington deemed this secondary data analysis on deidentified data “Not Human Subjects Research” (ref. 16421).

## RESULTS

### Sample characteristics

Of the 5,059 people enrolled in HAALSI, n=2,345 (46%) were men, and n=2714 (54%) were women (Table 1). In both men and women, the groups with five or more living children were the largest (51% [median: 5, IQR: 3-7] and 47% [median: 4 IQR:3-6, respectively). There were no major differences by sex, though men were more highly educated, more likely to be literate, and had higher mean cognitive performance. (Table 1, Figure 2)

**Table 1.** Sample characteristics by sex/gender from HAALSI*, Agincourt, South Africa, 2014/2015.

| Characteristic | Overall<br>N = 5,059<sup>1</sup> | Men<br>n = 2,345<sup>1</sup> | Women<br>n= 2,714<sup>1</sup> | p-value<sup>2</sup> |
| --- | --- | --- | --- | --- |
| **Number of living children** |  |  |  | <0.001 |
| 0 children | 311 (6.2%) | 187 (8.0%) | 124 (4.6%) |  |
| 1 to 2 children | 863 (17%) | 387 (17%) | 476 (18%) |  |
| 3 to 4 children | 1,379 (28%) | 560 (24%) | 819 (31%) |  |
| 5 or more children | 2,456 (49%) | 1,191 (51%) | 1,265 (47%) |  |
| Unknown | 50 | 20 | 30 |  |
| **Age** | 62 (13) | 62 (13) | 62 (13) | 0.6 |
| **Self-rated childhood health** |  |  |  | 0.063 |
| Very good | 3,501 (69%) | 1,585 (68%) | 1,916 (71%) |  |
| Good | 933 (18%) | 449 (19%) | 484 (18%) |  |
| Moderate | 296 (5.9%) | 157 (6.7%) | 139 (5.1%) |  |
| Bad | 164 (3.2%) | 81 (3.5%) | 83 (3.1%) |  |
| Very bad | 161 (3.2%) | 72 (3.1%) | 89 (3.3%) |  |
| Unknown | 4 | 1 | 3 |  |
| **Birth Country** |  |  |  | 0.11 |
| South Africa | 3,528 (70%) | 1,663 (71%) | 1,865 (69%) |  |
| Mozambique/other | 1,526 (30%) | 682 (29%) | 844 (31%) |  |
| Unknown | 5 | 0 | 5 |  |
| **Father's job skill level** |  |  |  | <0.001 |
| Level 1 | 1,446 (32%) | 649 (30%) | 797 (34%) |  |
| Level 2 | 2,187 (49%) | 1,118 (52%) | 1,069 (46%) |  |
| Level 3 | 151 (3.4%) | 68 (3.2%) | 83 (3.5%) |  |
| Level 4 | 141 (3.1%) | 64 (3.0%) | 77 (3.3%) |  |
| Other | 575 (13%) | 253 (12%) | 322 (14%) |  |
| Unknown | 559 | 193 | 366 |  |
| **Education** |  |  |  | <0.001 |
| No formal education | 2,306 (46%) | 957 (41%) | 1,349 (50%) |  |
| Some formal education | 2,736 (54%) | 1,381 (59%) | 1,355 (50%) |  |
| Unknown | 17 | 7 | 10 |  |
| **Literacy** |  |  |  | <0.001 |
| Illiterate | 2,108 (42%) | 764 (33%) | 1,344 (50%) |  |
| Literate | 2,948 (58%) | 1,580 (67%) | 1,368 (50%) |  |
| Unknown | 3 | 1 | 2 |  |
| **Cognitive status** |  |  |  | 0.003 |
| Cognitively normal | 4,603 (92%) | 2,158 (93%) | 2,445 (91%) |  |
| Cognitively impaired | 416 (8.3%) | 163 (7.0%) | 253 (9.4%) |  |
| Unknown | 40 | 24 | 16 |  |
| **Standardized cognitive score** | 0.00 (1.00) | 0.07 (0.99) | -0.06 (1.01) | <0.001 |
| Unknown | 156 | 80 | 76 |  |

<sup>1</sup>n (%); Mean (SD), <sup>2</sup>Pearson's Chi-squared test; Wilcoxon rank sum test
\*HAALSI: Health and Aging in Africa: A Longitudinal Study in South Africa

**Figure 2.**
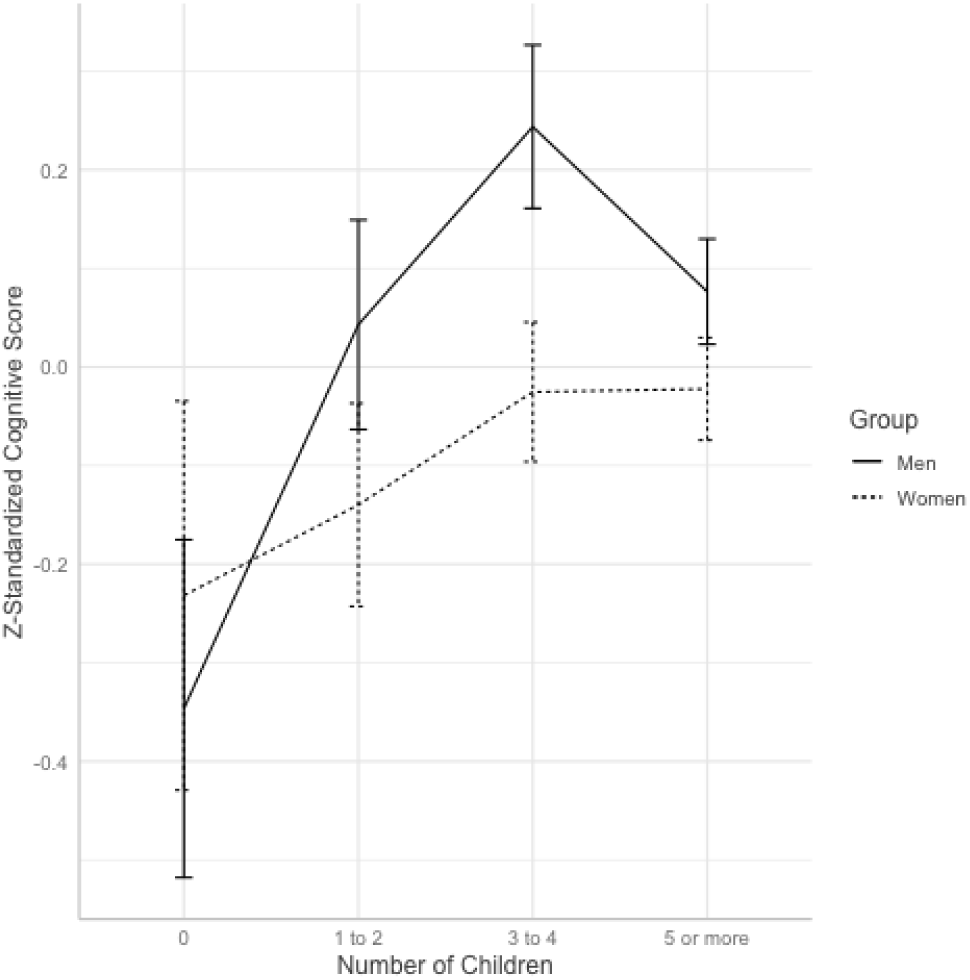
Mean Z-standardized cognitive score by categorical number of children for men and women in HAALSI, Agincourt, South Africa, 2014/2015. Error bars display the standard error.

Among men, 187 (8%) had no living children, 387 (17%) had 1 to 2 living children, 560 (24%) had 3 or 4 living children, and 1,191 (51%) had five or more living children (Table 1). Age increased with the number of living children, and literacy and education were highest among those with 1-2 and 3-4 children (Supplemental Table 1). There was an inverse U-shaped relationship between number of children and cognitive function, with the lowest mean cognitive scores and the greatest prevalence of cognitive impairment observed in those with 0 children (Supplemental Table 1, Figure 2).

For women, 124 (5%) had no living children, 476 (18%) had 1 to 2 living children, 819 (31%) had 3 to 4 living children, and 1,265 (47%) had five or more children (Table 1). The mean age was similar across groups (Supplemental Table 2). Mean cognitive scores increased as the number of children increased, with the lowest mean cognitive scores and the greatest prevalence of cognitive impairment observed in the group with no children (Supplemental Table 2, Figure 2).

### Cognitive function

We found that, among men, all number of children categories were associated with higher cognitive function relative to having no children (Table 2). In the fully adjusted model, compared to men with 0 living children, we found men with 1 to 2 living children had a mean cognitive z-score that was 0.29 (95% CI: 0.13 – 0.45) standard deviations (SDs) higher, men with 3 to 4 living children had scores 0.50 (95% CI: 0.34 – 0.65) SDs higher, and men with five or more children had scores 0.48 (95% CI: 0.33 – 0.63) SDs higher. We also found that the group 3 to 4 living children had scores 0.21 SDs higher than those with 1 to 2 living children (95% CI: 0.09 – 0.33). Men with five or more living children had scores 0.19 SDs higher than those with 1-2 children (95% CI: 0.09 – 0.30) (p=0.0014). We found no difference between those with 3 to 4 children and five or more living children.

**Table 2.** Model estimates from linear regression and modified Poisson regression for men, including only living children from HAALSI*, Agincourt, South Africa, 2014/2015.

| **Linear regression predicting z-standardized latent cognitive function scores and 95% confidence intervals (CIs)** |  |  |  |
| --- | --- | --- | --- |
|  | **Model 1**<br>**(Unadjusted)** | **Model 2**<br>**(Partial adjustment)<sup>1</sup>** | **Model 3**<br>**(Full adjustment)<sup>2</sup>** |
| *Predictors* | *Estimates*<br>*(CIs)* | *Estimates*<br>*(CIs)* | *Estimates*<br>*(CIs)* |
| 1 to 2 children | **0.39**<br>**(0.21 – 0.57)** | **0.35**<br>**(0.19 – 0.51)** | **0.29**<br>**(0.13 – 0.45)** |
| 3 to 4 children | **0.59**<br>**(0.42 – 0.76)** | **0.54**<br>**(0.38 – 0.69)** | **0.50**<br>**(0.34 – 0.65)** |
| 5 or more children | **0.42**<br>**(0.26 – 0.58)** | **0.51**<br>**(0.37 – 0.66)** | **0.48**<br>**(0.33 – 0.63)** |
| Observations | 2246 | 2242 | 2061 |
| R<sup>2</sup> / R<sup>2</sup> adjusted | 0.021 / 0.019 | 0.235 / 0.233 | 0.274 / 0.269 |
| AIC | 6272.791 | 5710.902 | 5121.422 |

| **Prevalence ratios and 95% confidence intervals (CIs) for cognitive impairment from modified Poisson regression** |  |  |  |
| --- | --- | --- | --- |
|  | *Prevalence Ratios*<br>*(CIs)* | *Prevalence Ratios*<br>*(CIs)* | *Prevalence Ratios*<br>*(CIs)* |
| 1 to 2 children | **0.41**<br>**(0.27 – 0.64)** | **0.47**<br>**(0.31 – 0.72)** | **0.48**<br>**(0.31 – 0.75)** |
| 3 to 4 children | **0.21**<br>**(0.13 – 0.34)** | **0.28**<br>**(0.17 – 0.44)** | **0.21**<br>**(0.12 – 0.37)** |
| 5 or more children | **0.28**<br>**(0.19 – 0.40)** | **0.30**<br>**(0.21 – 0.43)** | **0.28**<br>**(0.19 – 0.41)** |
| Observations | 2301 | 2294 | 2106 |
| R<sup>2</sup> Nagelkerke | 0.058 | 0.281 | 0.312 |
| AIC | 1128.836 | 953.671 | 810.846 |

<sup>1</sup>Adjusted for age, education (reference: no education), and literacy (reference: cannot read or write)
<sup>2</sup>Adjusted for age, education (reference: no education), literacy (reference: cannot read or write), self-rated childhood health (reference: very good), birth country (reference: South Africa), father's job skill level (reference: level 1)
\*HAALSI: Health and Aging in Africa: A Longitudinal Study in South Africa

### Cognitive impairment

Men with any number of living children were less likely to be cognitively impaired than those with no children (Table 2). In the fully adjusted model, the prevalence of cognitive impairment among men with 1-2 living children was 52% lower (aPR [95% CI] 0.48 [0.31 - 0.75]) than that of men with no living children. Among men with 3-4 living children, the prevalence was 79% lower (aPR[95% CI] 0.21 [0.12 - 0.37]), and among men with ≥5 living children, the prevalence was 72% lower (aPR[95% CI] 0.28 [0.19 – 0.41]) than that of men with no children. Additionally, we found that the prevalence of cognitive impairment among those with 3 to 4 children was 56% lower than those with 1 to 2 children (aPR[95% CI] 0.44 [0.25 - 0.78]), while the prevalence among those with five or more children was 42% lower than those with 1 to 2 children (aPR[95% CI] 0.58 [0.38 - 0.87]). There was no difference in the prevalence of cognitive impairment between men with five or more children and those with 3 to 4 children (aPR[95% CI] 1.32 [0.77 - 2.27]).

Similarly, women with any number of living children were less likely to be cognitively impaired than those without children (Table 3). The prevalence of cognitive impairment among women with 1-2 children was 48% lower (aPR [95%CI] 0.52 [0.31 - 0.88]) compared to those with no living children, the prevalence among women with 3 to 4 children was 45% lower (aPR [95%CI] 0.55 [0.33 - 0.91]) that of women with no children, and women with five or more living children had a prevalence of cognitive impairment that was 59% lower (aPR [95%CI) 0.41 [0.25 - 0.69]) that of women with no living children (Table 3). We found no additional between-group differences.

**Table 3.** Model estimates from linear regression and modified Poisson regression for women, including only living children from HAALSI*, Agincourt, South Africa, 2014/2015.

| **Linear regression predicting z-standardized latent cognitive function scores and 95% confidence intervals (CIs)** |  |  |  |
| --- | --- | --- | --- |
|  | **Model 1 (Unadjusted)** | **Model 2 (Partial adjustment)<sup>1</sup>** | **Model 3 (Full adjustment)<sup>2</sup>** |
| *Predictors* | *Estimates (CIs)* | *Estimates (CIs)* | *Estimates (CIs)* |
| 1 to 2 children | 0.09<br>(-0.11 – 0.30) | 0.09<br>(-0.08 – 0.26) | 0.11<br>(-0.07 – 0.29) |
| 3 to 4 children | **0.21**<br>**(0.01 – 0.40)** | 0.07<br>(-0.09 – 0.23) | 0.11<br>(-0.06 – 0.28) |
| 5 or more children | **0.21**<br>**(0.02 – 0.40)** | 0.13<br>(-0.02 – 0.29) | **0.17**<br>**(0.01 – 0.34)** |
| Observations | 2610 | 2604 | 2263 |
| R<sup>2</sup> / R<sup>2</sup> adjusted | 0.003 / 0.002 | 0.343 / 0.341 | 0.349 / 0.345 |
| AIC | 7412.337 | 6321.508 | 5444.582 |
| **Prevalence ratios (PR) and 95% confidence intervals (CIs) for cognitive impairment from modified Poisson regression** |  |  |  |
|  | *Prevalence Ratios (CIs)* | *Prevalence Ratios (CIs)* | *Prevalence Ratios (CIs)* |
| 1 to 2 children | 0.82<br>(0.51 – 1.32) | 0.73<br>(0.47 – 1.13) | **0.52**<br>**(0.31 – 0.88)** |
| 3 to 4 children | **0.60**<br>**(0.38 – 0.963)** | 0.72<br>(0.47 – 1.11) | **0.55**<br>**(0.33 – 0.91)** |
| 5 or more children | **0.44**<br>**(0.28 – 0.70)** | **0.57**<br>**(0.37 – 0.89)** | **0.41**<br>**(0.25 – 0.69)** |
| Observations | 2668 | 2660 | 2300 |
| R<sup>2</sup> Nagelkerke | 0.020 | 0.375 | 0.399 |
| AIC | 1625.376 | 1267.874 | 960.872 |

<sup>1</sup>Adjusted for age, education (reference: no education), and literacy (reference: cannot read or write)
<sup>2</sup>Adjusted for age, education (reference: no education), literacy (reference: cannot read or write), self-rated childhood health (reference: very good), birth country (reference: South Africa), father's job skill level (reference: level 1)
\*HAALSI: Health and Aging in Africa: A Longitudinal Study in South Africa

### Sensitivity analyses

In a sensitivity analysis to check the robustness of decisions around coding our exposure, we found that results for both men and women remained similar when we included deceased children in the definition of the categorical total number of children (supplemental tables 3 and 4). We found men with any number of total children to have higher cognitive scores and a lower prevalence of cognitive impairment than men without children with effect sizes comparable to the primary analysis (supplemental table 3). In women, we found the prevalence of cognitive impairment was lower in all groups with any number of total children compared to women without children with effect sizes comparable to the primary analysis (supplemental table 4).

To explore the potential for non-linear relationships between number of children and cognitive scores, we used the living number of children as a continuous exposure with a quadratic term in the linear regression models. For men, we found evidence for a non-linear relationship (upside down U shaped), with a statistically significant quadratic term [adjusted B (95% CI): −2.77 (−4.55, −0.98)] (supplemental table 5 and supplemental figure 1). Among women, no evidence for a non-linear relationship was observed (supplemental table 6 and supplemental figure 2).

## DISCUSSION

In this population-based sample of aging adults from rural South Africa during 2014/2015, we found that having living children was associated with greater cognitive functioning and lower prevalence of cognitive impairment. This association remained after adjusting for known risk and socioeconomic factors. In the adjusted models, we found that, among men, there was a relatively large protective association with having any living children for both cognitive function and cognitive impairment. For women, having any number of children was associated with a lower prevalence of cognitive impairment, but only the group with 5 or more children showed a relatively small effect size, indicating better cognitive performance, compared to those with no living children.

Prior work assessing the relationship between number of children and cognitive function has been varied. For men, our results were consistent with selected prior research,(23–26) showing a U-shaped association between number of children and prevalence of cognitive impairment and an inverse U-shaped relationship between number of children and cognitive function that remained after adjustment. In other words, the prevalence of cognitive impairment is lowest, and cognitive function is highest in those men with a moderate number of children. For women, our cognitive function results were inconsistent with prior research, which showed either decreasing cognitive performance as the number of children increased(27–37) or an inverse u-shaped relationship, with greater cognitive performance in those with a moderate number of children.(22–26) We found a positive association for cognitive function; women with 5 or more living children had greater cognitive function scores than those without children. The prevalence of cognitive impairment was lower for women with any number of children relative to those with 0 children. We observed that the women with five or more children had the lowest prevalence ratio, indicating that the prevalence of cognitive impairment was 59% lower in this group compared to women with 0 children.

We expected that the experience of having and raising children would contribute to cognitive function and impairment in positive and negative ways across the lifespan (Figure 1). As such, we were particularly interested in examining the total effect of having living children rather than attempting to isolate a single factor. Some of these factors were more likely to be experienced by the pregnant parent or the parent who participated most in childrearing. The biological experience of pregnancy and childbirth is unique to the pregnant parent and, therefore, may explain some of the sex/gender differences that we have observed in this study. In South Africa, previous work has found that mothers perform many day-to-day tasks related to childrearing, with various barriers preventing men from being active participants or being acknowledged as active participants. However, fathers are often expected to and may contribute financially.(53–58) In this population, men may reap more of the cognitive benefits of having children without experiencing the cognitive costs. For example, men and women in rural South Africa may both receive the cognitive benefits of having and raising children, but women are performing more of the tasks that result in cognitive costs, such as pregnancy and sleep loss.

Pregnancy may contribute to later-life cognitive dysfunction, resulting in physiological changes associated with reduced cognitive function. These include reductions in high-density lipoprotein and changes in glucose metabolism, which persist into later life and are associated with later-life cognition.(10,11,59–61) There are hormonal changes following pregnancy and the birth of a child. For women, estrogen and estradiol levels vary through the lifespan, and these variations can be associated with reproductive life history.(8,9,62,63) For example, estrogens, which tend to have neuroprotective effects, increase during pregnancy but then drop following birth.(8,62–64) Mothers also experience postpartum cognitive deficits, which can persist into later life.(65–67) While men do not experience pregnancy and childbirth, they experience hormonal changes after becoming a father. Testosterone, which is also neuroprotective, decreases after becoming a father, especially when men actively participate in childrearing. (6,68) Men in this population may not participate in as many childrearing activities, so their testosterone levels remain elevated, preserving the neuroprotective effects, though this has not been confirmed. While both mothers and fathers undergo physiological/hormonal changes upon becoming a parent, the maternal experience may have greater negative cognitive effects, partially explaining the sex/gender differences in our results.

In addition to the biological effects of having a child, parents, particularly those heavily involved in childrearing, experience different social and behavioral influences than nonparents and parents not involved in childrearing. Involved parents sleep less, have more sleep disturbances, and experience more life-long stresses. Sleep disturbances in mid-life are associated with worse later-life cognition,(13,14,69–71) and stress from childrearing may be related to worse later-life cognition.(14–16,72) If women in this population assume most childrearing tasks, they may be differentially exposed to these factors, negatively influencing later-life cognitive performance.

Not all the factors involved in parenting negatively impact cognitive performance. Having and raising children also offers cognitive stimulation, potentially building cognitive reserve throughout the childrearing period.(73,74) Additionally, providing financial support through employment or other means may also be associated with increased cognitive reserve.(75) Fathers are expected to and do contribute financially to their children rather than participate in routine childrearing tasks, which could explain some of the sex/gender differences in our findings. Alternatively, men with higher cognition may be more likely to have children and show better mid- to later-life cognitive performance. Adult children may also provide social support and engagement in later life, though the impact on cognition is mixed.(15,17–19) On the other hand, being childless has been shown to have significant social and cultural consequences, particularly in low-resource areas.(76) Some childrearing activities substantially positively influence cognition, resulting in a potential net positive association between number of children and cognitive performance.

In this population, we found that having children was positively associated with cognition, though the effect was smaller in magnitude for women. As a result of these findings, it would be beneficial to explore targeted programming or interventions that could be adopted to help women reap similar cognitive benefits from having children as men. These interventions could include promoting programs for fathers to share the burden of childrearing tasks or providing additional support to mothers during these times.

This study has limitations. First, our exposure variable is limited by its self-reported nature. The survey questions do not differentiate between biological and nonbiological children. Since many of the pathways described above, particularly for women, influence cognition through the biological mechanism of having children, some participants may have their exposure misclassified if reporting on nonbiological children. If pregnancy indeed does have a considerable influence on cognition, the misclassification could bias the true relationship between childbearing and cognition.

Additionally, our main exposure includes living children only, potentially resulting in biased results. However, in our sensitivity analyses, we found that the magnitude and direction of the effect were similar when we included both living and deceased children in the exposure. Additionally, the cross-sectional nature of our analysis does not allow us to establish temporality between exposure and outcome or assess longitudinal rates of cognitive decline. While having children generally occurs before later-life cognitive decline or impairment, our results may be confounded by early-life factors influencing both parity and later-life cognition.(4,77) Finally, the measure of cognition used in this study is somewhat crude; future waves have utilized additional cognitive assessments to create a more robust picture of cognition. Future steps include exploring these relationships through longitudinal analyses that can assess rates of decline between the groups.

Strengths of this study include its large, population-representative sample of older adults in a rural South African setting. This large sample allowed us to observe potentially small effect sizes that may not have been evident in a smaller sample. Also, we chose to include men in our analyses. Much of the previous work examining the relationship between parity and later-life cognition focuses solely on women/mothers.(21,22,27,29–33,35–37) By including men, we can speculate about some differential associations between the biological and social aspects of parenting. As men and women have different biological and social roles in parenting and childrearing, stratifying the analysis allowed us to begin to distinguish between these influences.

In conclusion, we found that having living children was associated with greater cognitive functioning and a lower prevalence of cognitive impairment among older men and women in rural South Africa, with some evidence that there were larger effect sizes in men. Many factors, including biological and social factors, could influence the association between number of living children and mid-to-late life cognitive performance, and future work should seek to unpack these potential influences. The observed sex/gender difference may be due to differential exposures within the biological and social roles involved in childbearing and rearing. Providing support to parents may help women and men both receive cognitive benefits from having and raising children in later life.

## Data Availability Statement

The datasets analyzed for this study can be accessed at the following link: https://haalsi.org/data.

