## Supplemental Material for "Number of children and mid- to later-life cognitive function and cognitive impairment in rural South Africa: Evidence from “Health and Aging in Africa: A Longitudinal Study of an INDEPTH Community in South Africa” (HAALSI)"

**Supplemental Table 1. Sample Characteristics for Men by categorical number of children from HAALSI, Agincourt, South Africa, 2014/2015**

| Characteristic | 0 children  N = 187^1^ | 1 to 2 children  N = 387^1^ | 3 to 4 children  N = 560^1^ | 5 or more children  N = 1,191^1^ | p-value^2^ |
| --- | --- | --- | --- | --- | --- |
| Age | 55 (13) | 58 (13) | 60 (13) | 65 (12) | <0.001 |
| Self-rated childhood health |  |  |  |  | 0.017 |
| Very Good | 109 (58%) | 260 (67%) | 385 (69%) | 820 (69%) |  |
| Good | 42 (22%) | 72 (19%) | 104 (19%) | 224 (19%) |  |
| Moderate | 19 (10%) | 28 (7.2%) | 42 (7.5%) | 67 (5.6%) |  |
| Bad | 11 (5.9%) | 10 (2.6%) | 10 (1.8%) | 50 (4.2%) |  |
| Very bad | 6 (3.2%) | 17 (4.4%) | 19 (3.4%) | 29 (2.4%) |  |
| Unknown | 0 | 0 | 0 | 1 |  |
| Birth Country |  |  |  |  | 0.051 |
| South Africa | 137 (73%) | 283 (73%) | 415 (74%) | 815 (68%) |  |
| Mozambique/other | 50 (27%) | 104 (27%) | 145 (26%) | 376 (32%) |  |
| Father’s job skill level |  |  |  |  | 0.10 |
| Level 1 | 70 (40%) | 115 (32%) | 161 (32%) | 298 (27%) |  |
| Level 2 | 73 (42%) | 186 (51%) | 261 (51%) | 586 (54%) |  |
| Level 3 | 3 (1.7%) | 7 (1.9%) | 16 (3.1%) | 42 (3.9%) |  |
| Level 4 | 5 (2.9%) | 11 (3.0%) | 15 (2.9%) | 33 (3.0%) |  |
| Other | 22 (13%) | 46 (13%) | 58 (11%) | 125 (12%) |  |
| Unknown | 14 | 22 | 49 | 107 |  |
| Education |  |  |  |  | <0.001 |
| No formal education | 87 (47%) | 152 (39%) | 191 (34%) | 515 (43%) |  |
| Some formal education | 99 (53%) | 234 (61%) | 369 (66%) | 671 (57%) |  |
| Unknown | 1 | 1 | 0 | 5 |  |
| Literacy |  |  |  |  | <0.001 |
| Illiterate | 95 (51%) | 132 (34%) | 149 (27%) | 377 (32%) |  |
| Literate | 92 (49%) | 255 (66%) | 410 (73%) | 814 (68%) |  |
| Unknown | 0 | 0 | 1 | 0 |  |
| Cognitive Status |  |  |  |  | <0.001 |
| Cognitively Normal | 147 (80%) | 354 (92%) | 527 (96%) | 1,115 (94%) |  |
| Cognitively Impaired | 37 (20%) | 32 (8.3%) | 23 (4.2%) | 66 (5.6%) |  |
| Unknown | 3 | 1 | 10 | 10 |  |
| Standardized cognition | -0.35 (1.11) | 0.04 (1.04) | 0.24 (0.98) | 0.08 (0.93) | <0.001 |
| Unknown | 24 | 15 | 15 | 25 |  |
| ^1^n (%); Mean (SD),  ^2^Pearson's Chi-squared test; Kruskal-Wallis rank sum test | | | | | |

**Supplemental Table 2. Sample Characteristics for Women by categorical number of children from HAALSI, Agincourt, South Africa, 2014/2015**

| Characteristic | 0 children  N = 124^1^ | 1 to 2 children  N = 476^1^ | 3 to 4 children  N = 819^1^ | 5 or more children  N = 1,265^1^ | p-value^2^ |
| --- | --- | --- | --- | --- | --- |
| Age | 63 (14) | 64 (15) | 61 (14) | 61 (12) | 0.002 |
| Self-rated childhood health |  |  |  |  | 0.4 |
| Very Good | 85 (69%) | 334 (70%) | 575 (70%) | 903 (71%) |  |
| Good | 25 (20%) | 80 (17%) | 143 (17%) | 230 (18%) |  |
| Moderate | 9 (7.3%) | 24 (5.1%) | 42 (5.1%) | 62 (4.9%) |  |
| Bad | 4 (3.2%) | 22 (4.6%) | 24 (2.9%) | 32 (2.5%) |  |
| Very bad | 1 (0.8%) | 15 (3.2%) | 35 (4.3%) | 37 (2.9%) |  |
| Unknown | 0 | 1 | 0 | 1 |  |
| Birth Country |  |  |  |  | 0.2 |
| South Africa | 91 (74%) | 326 (68%) | 581 (71%) | 853 (68%) |  |
| Mozambique/other | 32 (26%) | 150 (32%) | 238 (29%) | 410 (32%) |  |
| Unknown | 1 | 0 | 0 | 2 |  |
| Father’s job skill level |  |  |  |  | 0.2 |
| Level 1 | 28 (28%) | 151 (36%) | 227 (32%) | 374 (34%) |  |
| Level 2 | 49 (49%) | 185 (44%) | 348 (49%) | 481 (44%) |  |
| Level 3 | 3 (3.0%) | 15 (3.6%) | 14 (2.0%) | 50 (4.6%) |  |
| Level 4 | 2 (2.0%) | 13 (3.1%) | 27 (3.8%) | 35 (3.2%) |  |
| Other | 18 (18%) | 55 (13%) | 100 (14%) | 147 (14%) |  |
| Unknown | 24 | 57 | 103 | 178 |  |
| Education |  |  |  |  | 0.003 |
| No formal education | 74 (60%) | 250 (53%) | 370 (45%) | 634 (50%) |  |
| Some formal education | 49 (40%) | 223 (47%) | 447 (55%) | 629 (50%) |  |
| Unknown | 1 | 3 | 2 | 2 |  |
| Literacy |  |  |  |  | 0.005 |
| Illiterate | 67 (54%) | 257 (54%) | 366 (45%) | 635 (50%) |  |
| Literate | 57 (46%) | 219 (46%) | 453 (55%) | 630 (50%) |  |
| Cognitive status |  |  |  |  | <0.001 |
| Cognitively Normal | 104 (85%) | 414 (87%) | 738 (91%) | 1,172 (93%) |  |
| Cognitively Impaired | 19 (15%) | 60 (13%) | 76 (9.3%) | 85 (6.8%) |  |
| Unknown | 1 | 2 | 5 | 8 |  |
| Standardized cognition | -0.23 (1.08) | -0.14 (1.12) | -0.03 (1.02) | -0.02 (0.93) | 0.028 |
| Unknown | 7 | 19 | 21 | 27 |  |
| ^1^n (%); Mean (SD),  ^2^ Pearson’s Chi-squared test; Kruskal-Wallis rank sum test; Fisher’s Exact Test for Count Data with simulated p-value (based on 1e+05 replicates) | | | | | |

| **Supplemental Table 3. Model estimates for men, including both living and deceased children from HAALSI, Agincourt, South Africa, 2014/2015** | | | |
| --- | --- | --- | --- |
| Linear regression predicting z-standardized latent cognitive function scores and 95% confidence intervals (CIs) | | | |
|  | **Model 1**  **(Unadjusted)** | **Model 2**  **(Partial adjustment)^1^** | **Model 3**  **(Full adjustment)^2^** |
| *Predictors* | *Estimates*  *(CIs)* | *Estimates*  *(CIs)* | *Estimates*  *(CIs)* |
| 1 to 2 children | **0.49 (0.30 – 0.68)** | **0.37 (0.20 – 0.54)** | **0.30 (0.13 – 0.47)** |
| 3 to 4 children | **0.63 (0.46 – 0.81)** | **0.51 (0.35 – 0.66)** | **0.46 (0.31 – 0.62)** |
| 5 or more children | **0.40 (0.24 – 0.56)** | **0.50 (0.36 – 0.65)** | **0.47 (0.32 – 0.62)** |
| Observations | 2242 | 2238 | 2059 |
| R^2^ / R^2^ adjusted | 0.023 / 0.022 | 0.233 / 0.231 | 0.272 / 0.266 |
| AIC | 6252.152 | 5703.411 | 5125.362 |
| Prevalence ratios (PR) and 95% confidence intervals (CIs) for cognitive impairment from modified Poisson regression | | | |
|  | **Model 1**  **(Unadjusted)** | **Model 2**  **(Partial adjustment)^1^** | **Model 3**  **(Full adjustment)^2^** |
| *Predictors* | *(95% CI)* | *PR*  *(95% CI)* | *PR*  *(95% CI)* |
| 1 to 2 children | **0.38 (0.22 – 0.64)** | **0.48 (0.27 – 0.82)** | **0.50 (0.27 – 0.87)** |
| 3 to 4 children | **0.21 (0.12 – 0.36)** | **0.32 (0.18 – 0.55)** | **0.30 (0.16 – 0.54)** |
| 5 or more children | **0.28 (0.19 – 0.42)** | **0.29 (0.20 – 0.44)** | **0.27 (0.17 – 0.42)** |
| Observations | 2296 | 2289 | 2103 |
| R^2^ Nagelkerke | 0.055 | 0.278 | 0.307 |
| AIC | 1119.819 | 946.217 | 809.159 |

| **Supplemental Table 4. Model estimates for women, including both living and deceased children from HAALSI, Agincourt, South Africa, 2014/2015** | | | |
| --- | --- | --- | --- |
|  | **Model 1**  **(Unadjusted)** | **Model 2**  **(Partial adjustment)^1^** | **Model 3**  **(Full adjustment)^2^** |
| *Predictors* | *Estimates* | *Estimates* | *Estimates* |
| 1 to 2 children | **0.43 (0.20 – 0.65)** | 0.17 (-0.02 – 0.35) | 0.19 (-0.01 – 0.39) |
| 3 to 4 children | **0.37 (0.17 – 0.57)** | 0.08 (-0.08 – 0.25) | 0.11 (-0.07 – 0.28) |
| 5 or more children | 0.10 (-0.09 – 0.29) | 0.11 (-0.05 – 0.26) | 0.14 (-0.02 – 0.31) |
| Observations | 2608 | 2602 | 2262 |
| R^2^ / R^2^ adjusted | 0.018 / 0.017 | 0.342 / 0.341 | 0.349 / 0.345 |
| AIC | 7369.091 | 6318.670 | 5443.872 |
| Prevalence ratios (PR) and 95% confidence intervals (CIs) for cognitive impairment | | | |
|  | **Model 1**  **(Unadjusted)** | **Model 2**  **(Partial adjustment)^1^** | **Model 3**  **(Full adjustment)^2^** |
| *Predictors* | *PR*  *(95% CI)* | *PR*  *(95% CI)* | *PR*  *(95% CI)* |
| 1 to 2 children | **0.45 (0.23 – 0.88)** | 0.67 (0.34 – 1.30) | **0.48 (0.21 – 1.07)** |
| 3 to 4 children | **0.39 (0.22 – 0.69)** | 0.61 (0.35 – 1.10) | **0.48 (0.25 – 0.96)** |
| 5 or more children | **0.63 (0.40 – 1.05)** | 0.67 (0.43 – 1.11) | **0.48 (0.28 – 0.88)** |
| Observations | 2666 | 2658 | 2299 |
| R^2^ Nagelkerke | 0.014 | 0.373 | 0.396 |
| AIC | 1630.499 | 1269.847 | 963.375 |
| **Supplemental Table 5. Model estimates for men, using living number of children as a continuous exposure with a quadratic term, HAALSI, Agincourt, South Africa, 2014/2015** | | | |
|  | **Standardized cognition** | **Standardized cognition** | **Standardized cognition** |
| *Predictors* | *Estimates* | *Estimates* | *Estimates* |
| Number of children | 1.58 (-0.39 – 3.55) | **4.78 (2.95 – 6.61)** | **4.53 (2.64 – 6.43)** |
| Number of children^2^ | **-2.01 (-3.97 – -0.05)** | **-2.34 (-4.08 – -0.60)** | **-2.77 (-4.55 – -0.98)** |
| Observations | 2246 | 2242 | 2061 |
| R^2^ / R^2^ adjusted | 0.003 / 0.002 | 0.226 / 0.224 | 0.266 / 0.261 |
| AIC | 6311.039 | 5733.965 | 5141.400 |

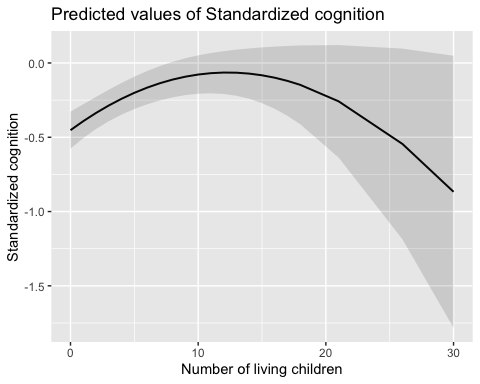

Supplemental figure 1: Predicted values of standardized cognition for each number of living children in men from HAALSI, Agincourt, South Africa 2014/15

| **Supplemental Table 6. Model estimates for women, using living number of children as a continuous exposure with a quadratic term, HAALSI, Agincourt, South Africa, 2014/2015** | | | |
| --- | --- | --- | --- |
|  | **Standardized cognition** | **Standardized cognition** | **Standardized cognition** |
| *Predictors* | *Estimates* | *Estimates* | *Estimates* |
| Number of children | **2.60 (0.61 – 4.59)** | **2.05 (0.42 – 3.67)** | **2.45 (0.72 – 4.18)** |
| Number of children^2^ | **-2.39 (-4.38 – -0.40)** | -0.25 (-1.87 – 1.37) | -0.47 (-2.21 – 1.27) |
| Observations | 2610 | 2604 | 2263 |
| R^2^ / R^2^ adjusted | 0.005 / 0.004 | 0.343 / 0.342 | 0.350 / 0.346 |
| AIC | 7407.417 | 6318.339 | 5440.947 |

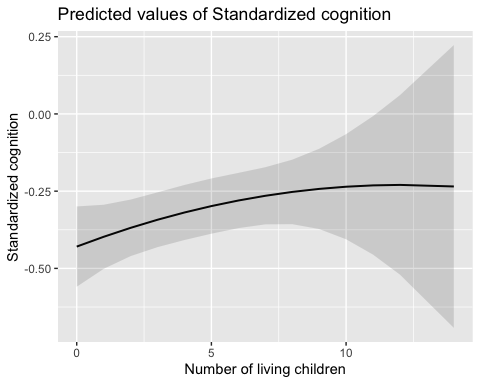

Supplemental figure 2: Predicted values of standardized cognition for each number of living children in women from HAALSI, Agincourt, South Africa 2014/15
